# Excess mortality during the COVID-19 pandemic: a geospatial and statistical analysis in Mogadishu, Somalia

**DOI:** 10.1101/2021.05.15.21256976

**Authors:** Abdihamid Warsame, Farah Bashiir, Terri Freemantle, Chris Williams, Yolanda Vazquez, Chris Reeve, Ahmed Aweis, Mohamed Ahmed, Francesco Checchi, Abdirisak Dalmar

## Abstract

**Background:** While the impact of the COVID-19 pandemic has been well documented in high-income countries, much less is known about its impact in Somalia where health systems are weak and vital registration is under developed.

**Methods:** We used remote sensing and geospatial analysis to quantify the number of burials from January 2017 to September 2020 in Mogadishu. We imputed missing grave counts using surface area data. Simple interpolation and a generalised additive mixed growth model were used to predict both actual and counterfactual burial rates by cemetery and across Mogadishu during the most likely period of COVID-19 excess mortality and to compute excess burials. We also undertook a qualitative survey of key informants to determine the drivers of COVID-19 excess mortality.

**Results:** Burial rates increased during the pandemic period with a ratio to pre-pandemic levels averaging 1.5-fold and peaking at 2.2-fold. When scaled to plausible range of baseline Crude Death Rates (CDR), excess death toll between January and September 2020 ranged between 3,200 and 11,800. When compared to burial records of the Barakaat Cemetery Committee our estimates were found to be lower.

**Conclusions:** Our study points to considerable under estimation of COVID-19 impact in Banadir and an overburdened public health system struggling to deal with the increasing severity of the epidemic in 2020.

## Introduction

As a result of three decades of protracted conflict and state fragility Somalia’s health system is considered very weak and reports some of the worst health indicators in the world(Directorate of National Statistics Federal Government of Somalia, 2020). The emergence of the COVID-19 pandemic has exacerbated the effects of ongoing humanitarian crises due to natural disasters and conflict. (UNDP, 2020; UNSC, 2020)

As of 6^th^ May 2021, the Somali government had reported 14,368 cumulative COVID-19 confirmed cases and 745 cumulative confirmed deaths due to COVID-19. (Ministry of Health Somalia, 2021)Although these figures are much lower than those of countries with robust public health infrastructure, there is concern that they do not fully reflect the reality of the epidemic in the country. As a result of the country’s very low COVID-19 testing capacity, the potential stigma associated with COVID-19, and limited accessibility to many parts of the country, the official data may not be an accurate representation of the true burden of COVID-19.(UNFPA Somalia, 2020; WHO, 2020) Investigative enquiries by news organizations and anecdotal reports point to a larger numbers of COVID-19 deaths than have been officially reported. (BBC News, 2020; Jason Burke and Abdalle Ahmed Mummin, 2020; Wariyaha Muqdisho, 2020) We attempted to shed light on the impact of COVID-19 in Mogadishu by estimating excess mortality using satellite imagery analysis as well as highlighting key contextual drivers using qualitative methods.

## Methods

### Study population and period

We estimated excess mortality attributable to COVID-19 and related social and health systems disruptions among people living in the Banadir region, which is almost entirely occupied by the rapidly expanding urban and peri-urban conurbation of Mogadishu. To estimate a baseline death rate, we defined a baseline period from 1 January 2017 to 31 December 2019, i.e. before widespread SARS-CoV-2 transmission in Mogadishu and an epidemic period from January to end September 2020, when we capped data collection. Under the assumption that all decedents in Banadir are buried in recognised cemeteries, we sought to identify and collect data on every cemetery that was ‘active’ (i.e. receiving new burials) at any point during the analysis period.

### Data Collection

#### Cemetery identification

An initial list of cemeteries in the Banadir region was identified via the analysis of open-source location data and satellite imagery(Google Earth, 2021, Google Maps, 2021, OpenStreetMap, 2021)supplemented by field observations of local researchers. Analysis excluded five private or family-owned cemeteries - all five appeared active upon visitation, but we were constrained by vegetation cover for Abaay Dhaxan and the inability to procure suitable satellite imagery for Jazeera and El Adde (Figure 1). A list of all cemeteries identified and visited is included in the supplementary file.

**Figure 1.**
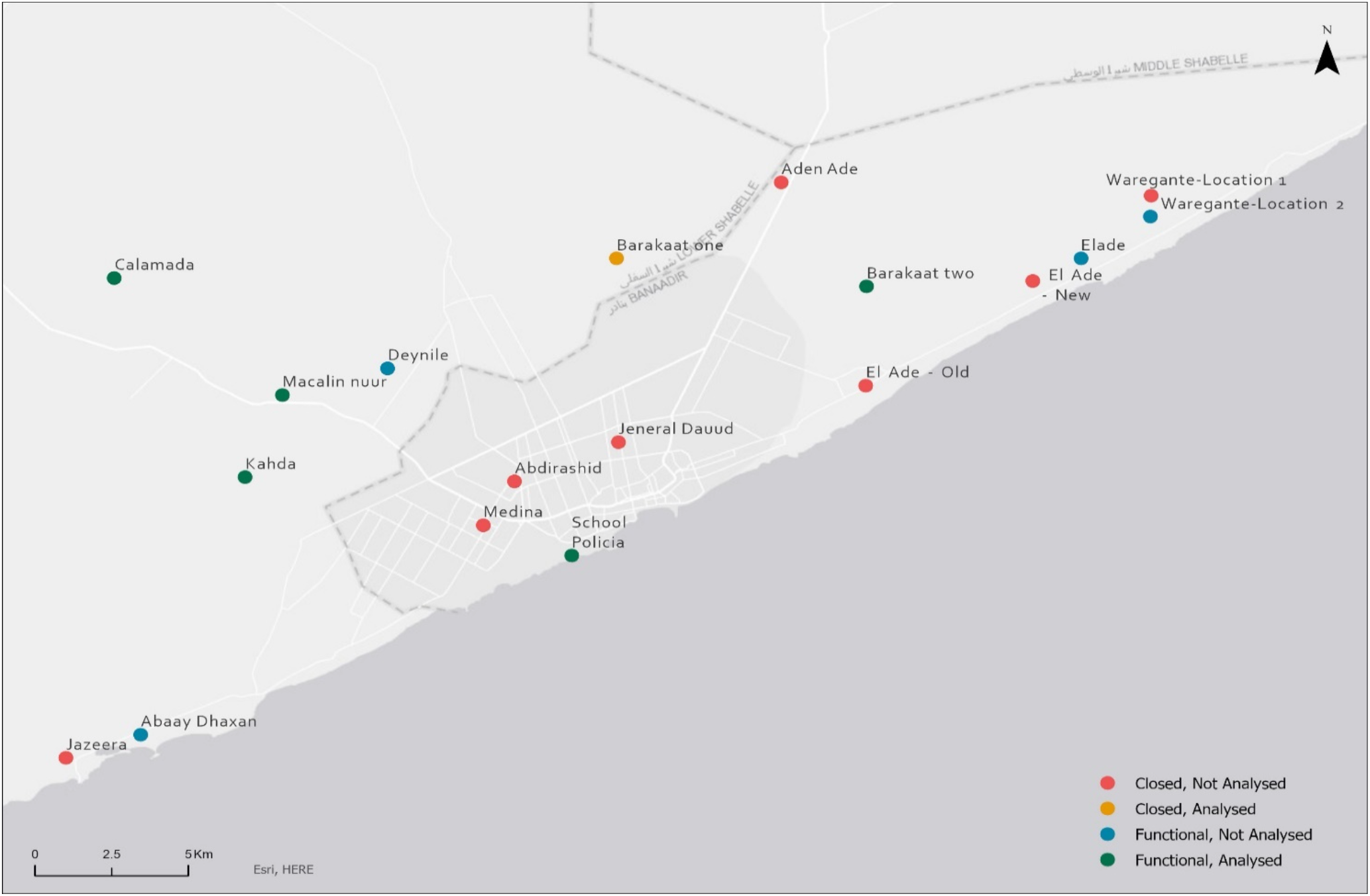
Location and status of cemeteries in Banadir.

#### Satellite imagery

The analysis relied on the availability of suitable archive satellite imagery acquired during the analysis period, with required pixel resolution and minimum visual obstruction. Where available, we sourced the most suitable available Very High Resolution (VHR) images for each cemetery during the analysis period, ensuring they were cloud-free, of high radiometric quality and most importantly, focussing on those with a spatial resolution between ∼31cm to ∼40cm per pixel. The higher resolution of 31cm was selected where available, as it provided the best clarity of features on the ground and ensured the most accurate identification of new burials. We used coarser resolution imagery (∼40cm to ∼50cm) where no higher resolution alternative was available, to investigate its uses, limitations and, at a minimum, measure surface area change. At least one satellite image per year was obtained for 2016 – 2019, if available; between January – October 2020, where available, we sourced monthly imagery. Images were purchased as Ortho Natural Colour via SecureWatch (Maxar, 2021) and were delivered pre-processed, corrected for illumination and geometric distortion, and pan-sharpened. To further improve the detection of individual burial plots, where beneficial some were enhanced using Edge Detection techniques (Sobel and/or Touzi filter(Centre national d’études spatiales, 2019)).

#### Other burial data sources

Separately, we obtained monthly reports by the Barakaat Cemetery Development Committee (BCDO) covering the periods April-June 2019 and January-April 2020.

#### Key Informant Interviews (KIIs)

A total of 20 key informant interviews were conducted with health authorities and professionals, gravediggers, religious leaders and other stakeholders to provide insights into community perception on COVID-19 mortality, community practice during the pandemic and challenges.

### Analysis

#### Population denominators

To compute burial rates per population, we computed population denominators for Banadir Region using two starting sources: an estimate by the WorldPop(Wordpop Gridded Population Estimate Datasets and Tools, 2020) projects for 2015and 2020 (assuming December as the month in which the estimate was centred). WorldPop redistributes existing countrywide estimates (e.g. through census exercises) across space using a predictive statistical model of population density per 100 m^2 pixels, built from various remotely-sensed variables including vegetation index and settlement pattern.(Linard C, Gilbert M, W. Snow R, Abdisalan M. Noor, 2012) Notably, for Somalia, the WorldPop project has developed methods to account for settlements for displaced persons. We forward-calculated population from 2015, and back-calculated from 2020, by using a natural growth rate of 30 per 1000 per year (assumed crude birth rate of 44 per 1000 per year minus crude death rate of 15 per 1000 per year, (the latter an average of existing surveys from Banadir Region: see Table 1), and by accounting for population movement in and out of the region over time, as reported by the United Nations High Commissioner for Refugees (UNHCR) Protection and Return Monitoring Project(UNHCR, n.d.). The two alternative base estimates yield ‘high’ and ‘low’ scenarios (Figure 2), which we carried into further estimation steps.

**Table 1.** Estimates (95% confidence intervals) from retrospective sample household surveys conducted in Banadir Region during 2013-2018 by humanitarian actors. Surveys investigated a retrospective period of 87-104 days. CDR = Crude death rate per 10,000 per day. Source: United Nations Food Security and Nutrition Analysis Unit for Somalia.

| livelihood | year | month | season | CDR | U5DR |
| --- | --- | --- | --- | --- | --- |
| displaced | 2013 | November | Deyr | 0.62 (0.3 to 1.3) | 0.5 (0.16 to 1.54) |
| displaced | 2015 | May | Gu | 0.63 (0.33 to 1.23) | 1.36 (0.72 to 2.54) |
| urban | 2015 | May | Gu | 0.54 (0.32 to 0.92) | 0.64 (0.27 to 1.49) |
| urban | 2015 | November | Deyr | 0.28 (0.15 to 0.53) | 0.23 (0.06 to 0.96) |
| agropastoralists | 2016 | May | Gu | 0.35 (0.16 to 0.73) | 1.02 (0.38 to 2.74) |
| displaced | 2016 | November | Deyr | 0.61 (0.34 to 1.11) | 0.74 (0.3 to 1.81) |
| displaced | 2017 | May | NA | 0.7 (0.36 to 1.33) | 1.28 (0.53 to 3.04) |
| urban | 2017 | May | NA | 0.68 (0.46 to 1.01) | 0.69 (0.32 to 1.49) |
| displaced | 2018 | March | NA | 0.42 (0.27 to 0.65) | 0.6 (0.25 to 1.41) |
| displaced | 2018 | May | Gu | 1.06 (0.73 to 1.54) | 2.56 (1.54 to 4.22) |
| urban | 2018 | June | Gu | 0.25 (0.11 to 0.54) | 0.34 (0.08 to 1.38) |
| displaced | 2018 | November | Deyr | 0.74 (0.48 to 1.12) | 1.21 (0.58 to 2.53) |
| urban | 2018 | November | Deyr | 0.15 (0.05 to 0.47) | 0.19 (0.03 to 1.46) |

**Figure 2.**
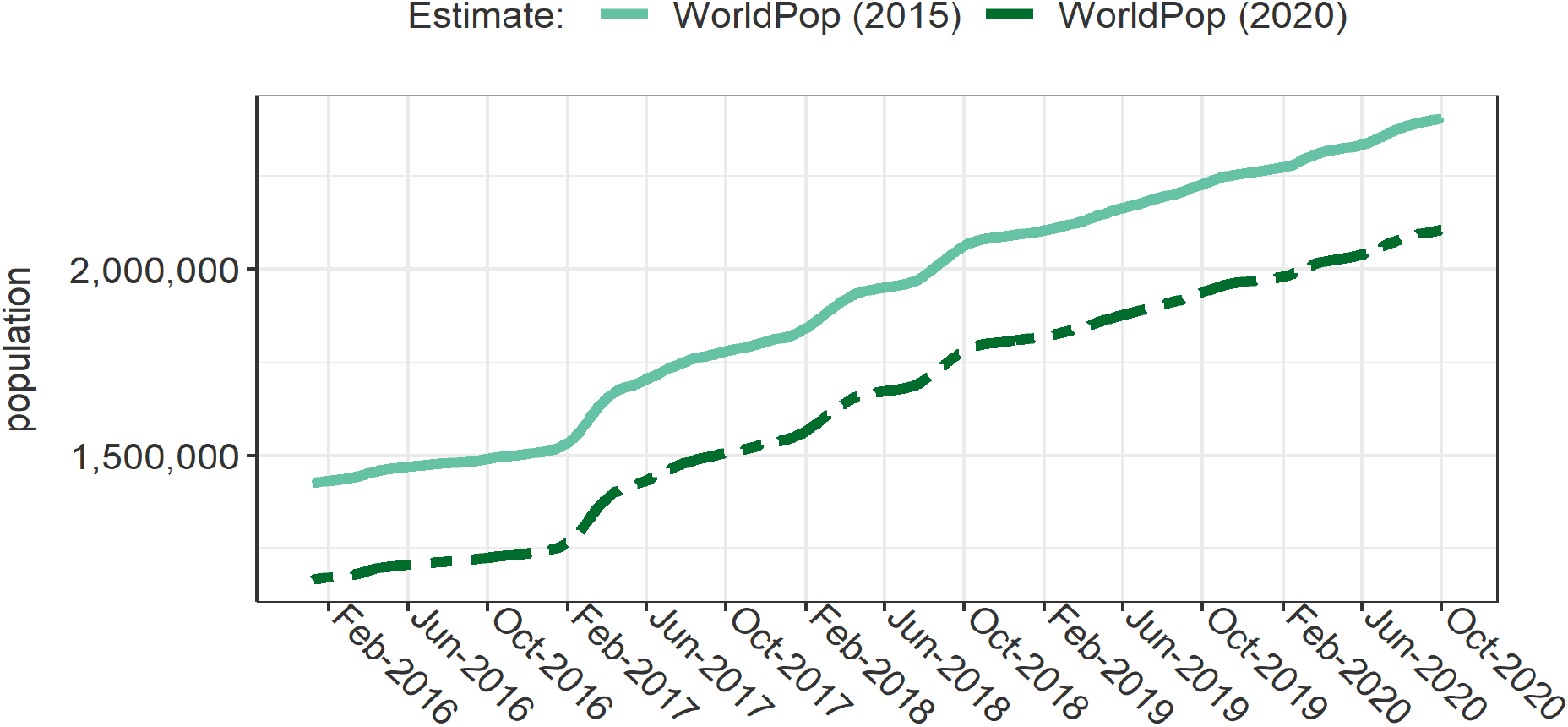
High and low estimates of the population over time in Banadir Region.

#### Imagery analysis

For the burial assessment, we combined manual and semi-automated image analysis approaches. The approaches applied were determined by the suitability of the input satellite imagery to a specified set of criteria – this included image quality, image resolution, cloud cover, the typology of the cemetery (formal vs informal arrangement, infilling vs new blocks) and the ability to identify individual burials plots by either a spectral or object-based image processing technique. Sites with imagery most compliant with the criteria (i.e. those in which the individual burial plots could be clearly observed) were analysed using a computer vision algorithm developed in Python to localise the georeferenced perimeter of regularly shaped, small, bright objects in optical satellite imagery. This was applied to extract individual burial plots and automatically compute a total count of features.

The output features were quality checked manually by an analyst, and in situations where the subsequent imagery was not of high enough quality to meet the criteria required for the computer vision, the features were used as a reference dataset during the manual annotation. Any sites classified as not meeting the pre-defined criteria were assessed manually by trained annotators. An analyst can annotate a satellite image by assigning semantic labels that represent certain features as objects. Whilst more time consuming, manual annotation results in lower uncertainty and higher precision when compared to automated and semi-automated annotation techniques. The use of expert annotators to create training label data was undertaken as the extraction of burial information from satellite imagery is a new and relatively untested approach(Besson et al., 2020). Furthermore, accurately labelled data is a prerequisite for the application of automated machine learning-based approaches that could facilitate the ability to undertake similar analyses at scale. Object-based image annotation(Cloudfactory, 2020) was undertaken to remotely locate and count the number of new individual burials within cemeteries when visible, for sites where individual plots were not identifiable, whether due to poor image quality or degradation of grave visibility through time – as the result of local climate and environment dynamics (e.g. erosion, sand migration) – the annotation was undertaken using a ‘bounding box’ to delineate the change in total burial area over time. Analysts manually tagged individual burials as ‘objects’ using point vectors.

### Statistical analysis

#### Inferring missing grave counts

Overall, 68 sequential satellite images were available across the six analysed cemeteries (mean 11.2 per cemetery). However, some images were constrained by cloud cover or poor data quality (e.g. inadequate resolution), such that, for a given time point, individual graves were not visible for a fraction or the totality of the cemetery, leaving the overall surface area of the cemetery as the sole robustly quantifiable variable. For an addition subset of images, poor image quality precluded a comprehensive grave count. Overall, we were able to perform an exhaustive grave count for 58.8% (40/68) of images. For 8 images (11.7%) we analysed only the visible area or a sample of 10 m^2^ quadrats selected randomly by overlaying a grid onto the cemetery: here, we extrapolated the ratio of graves per m^2^ observed within the sampled quadrats to approximate the total number of graves (for the first observation in a cemetery) or new graves since the previous image (for further time points). Since graves must increase with expanding cemetery surface areas, we imputed the number of graves in the remaining 20 (29.4%) images by fitting a generalised additive mixed model for location, size and shape (GAMLSS) of the count of new graves, with the natural log of new surface area as growth predictor, new surface area nested within the cemetery as a random effect and assuming a quasi-poisson distribution. We then used the model to predict the missing grave observations. Model output is shown in the supplementary file and all data and R scripts are available on Github (Checchi, 2021).

#### Estimating excess mortality

After linearly interpolating the time series of observed and imputed grave counts for each cemetery, we summed each time series to compute an overall trend for all cemeteries analysed, i.e. our estimate of burials across the Banadir region. We bound estimation to the period for which data were available for all cemeteries (1 January 2017 to 16 September 2020). We computed daily burial rates per 10,000 person-days for each alternative population estimate. After inspecting the resulting trend, we decided that the most reasonable, albeit crude, approach for defining a pre-COVID-19 baseline level of burials was to fit a smooth spline function to the trend in burial rate pre-2020 and extrapolate this forward into 2020 to compute a non-pandemic counterfactual.

Since the burial rate as estimated from satellite imagery appeared implausibly low, we used an indirect method for excess mortality estimation, informed by a range of plausible pre-pandemic crude death rate (CDR) values. Specifically, we scaled the estimated burial rate at 1 Jan 2017 to CDR values within the range 0.20 to 0.60 per 10,000 person-days: this reflects the range of results of rapid anthropometric and retrospective mortality surveys done by humanitarian actors between 2013 and 2018 in different communities of Banadir Region (Table 1).

We used each scaled time series of baseline CDR, in combination with the observed daily ratios of estimated to counterfactual burial rate during 2020 as well as population denominators, to estimate the actual and excess death rates across the Banadir Region, and the corresponding death tolls.

#### Qualitative Data Analysis

We transcribed and analysed the key informant interviews using thematic analysis (Bryman, 2012).

## Results

### Availability of Satellite Imagery

We included six cemeteries in analysis across Mogadishu and Banadir for which we were able to source imagery that met inclusion criteria (Table 2). The number of images varied among sites from 8 to 14. The earliest images were from January 2016 and the latest from October 2020. In total we tallied 18,616 burials across all sites during this period.

**Table 2.** Availability of imagery and characteristics of cemeteries included in the analysis.

| Cemetery | District | Typology | Status |  | N images | Imagery timespan |  | New graves during the period | Surface area (m<sup>2</sup>) |  |
| --- | --- | --- | --- | --- | --- | --- | --- | --- | --- | --- |
|  |  |  | 2016 | 2020 |  | Earliest | Latest |  | Start | End |
| Barakaat 1 | Yaqshid | New blocks and infilling | existed | closed | 14 | 20/02/2016 | 01/10/2020 | 5562 | 10,674 | 95,856 |
| Barakaat 2 | Heliwaa | New blocks and infilling | did not exist | open | 9 | 17/11/2019 | 25/09/2020 | 1992 | 0 | 18,391 |
| Calamada | Afgooye | New blocks | existed | open | 11 | 19/01/2016 | 16/09/2020 | 5161 | 15,574 | 41,155 |
| Iskool Bolisii | Xamarjajab | New blocks | existed | open | 8 | 02/11/2016 | 25/09/2020 | 720 | n/a | n/a |
| Kahda | Kaxda | Infilling | existed | open | 13 | 03/10/2016 | 01/10/2020 | 1325 | 5012 | 12,190 |
| Moallim Nuur | Deynile (Garasbaley) | Infilling | existed | open | 12 | 07/01/2016 | 25/09/2020 | 3856 | 72,335 | 93,442 |

Our analysis shows that cemeteries varied greatly in size with the largest, Barakaat One, with an estimated surface area of 95,856m^2^ and the smallest, Kahda, with 12,190 m^2^ by the end of the study period. In general, the largest burial sites were located in less densely populated areas and in many cases on the outskirts of Mogadishu.

Cemeteries also displayed a marked difference in expansion patterns. In the larger, professionally managed cemeteries, new burials occurred in new evenly spaced blocks at the edges of the cemetery thereby expanding the overall area of the cemetery. Other sites, presumably due to limitations in space, utilized ‘infilling’, whereby new burials were observed between existing burial rows (Figure 3).

**Figure 3.**
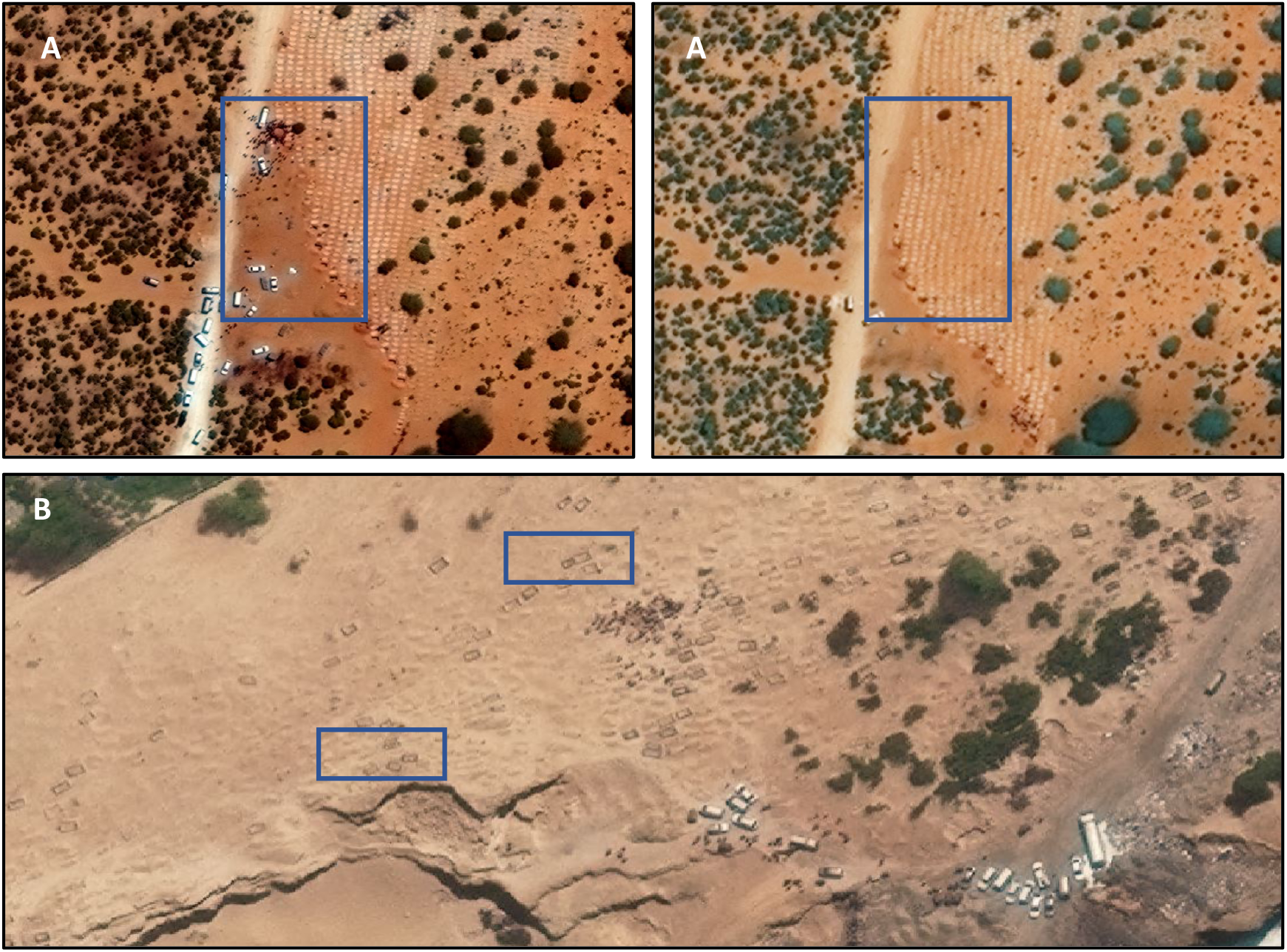
Sample of very high-resolution images from two cemeteries in Banadir, exemplifying the two typologies or burial pattern observed: (A) expansion into new ‘blocks’ and (B) ‘infilling’ within existing burial area Satellite imagery © Maxar Technologies 2021.

**Figure 4.**
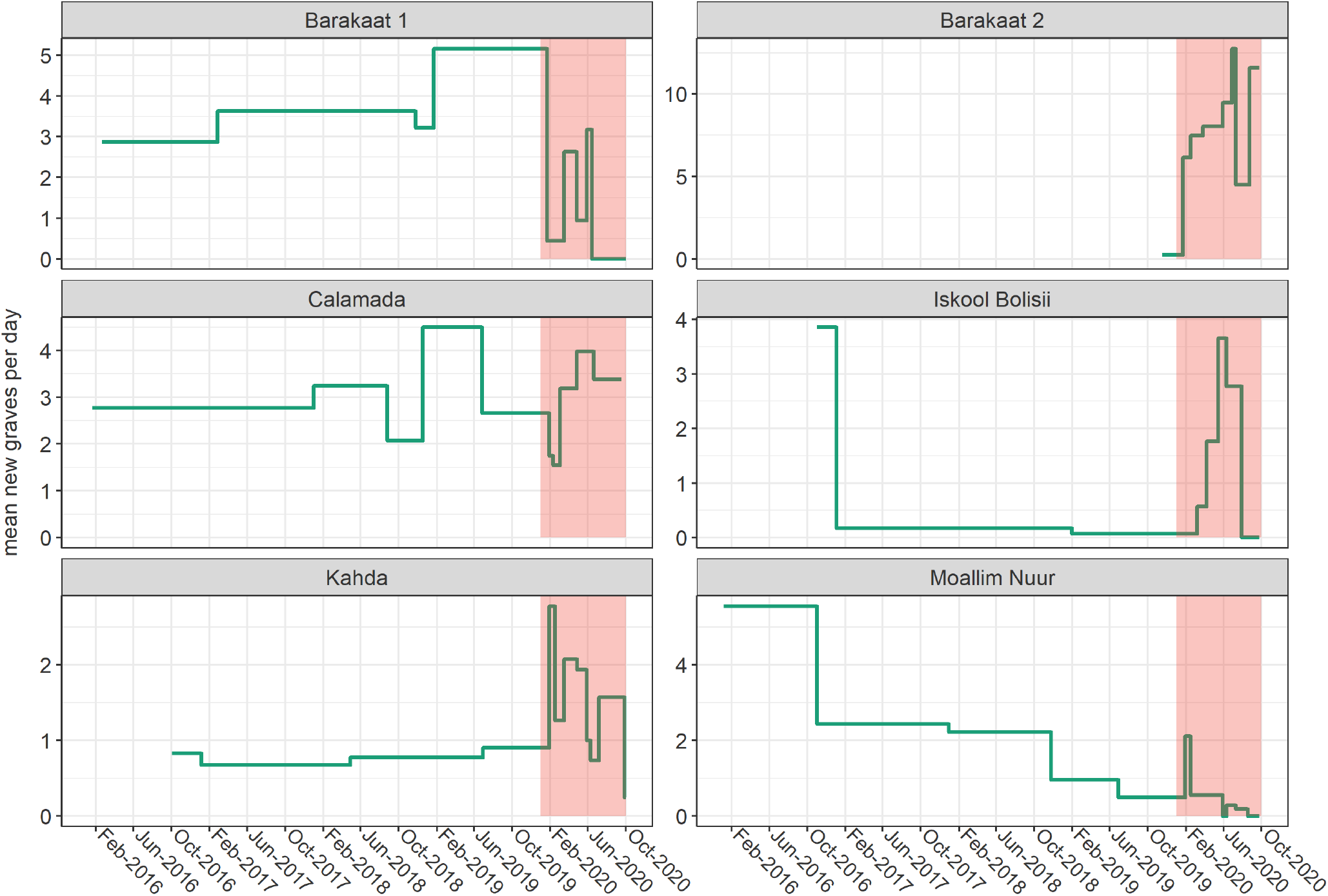
Evolution of burial rate over the analysis period, by cemetery. Each horizontal segment comprises the timespan between consecutive images. The assumed epidemic period (1 January 2020 onwards) is shaded in red. Note that the y-axis scales are cemetery-specific.

Burial rates across the analysed cemeteries demonstrated variability in the mean new graves observed per day during the analysis period. Some cemeteries displayed a relative increase in burials during the epidemic period (January 2020 onwards) with Iskool Bolisi and Barakaat 2 showing the most marked increase relative to baseline.

### Burial trends and excess mortality

Interpolated trends in daily burials, by cemetery, are shown in Figure 5. During the presumed baseline period (2017-2019), daily burials across the six cemeteries averaged about 10-12 (panel A). From January 2020, an increase in burials, peaking at >20 daily in June 2020, was evident. Note that trends in 2017-2019 are informed by only a few images (panel B), and as such may mask a more unstable pattern than that derived through our interpolation (i.e. unseen peaks and troughs).

**Figure 5.**
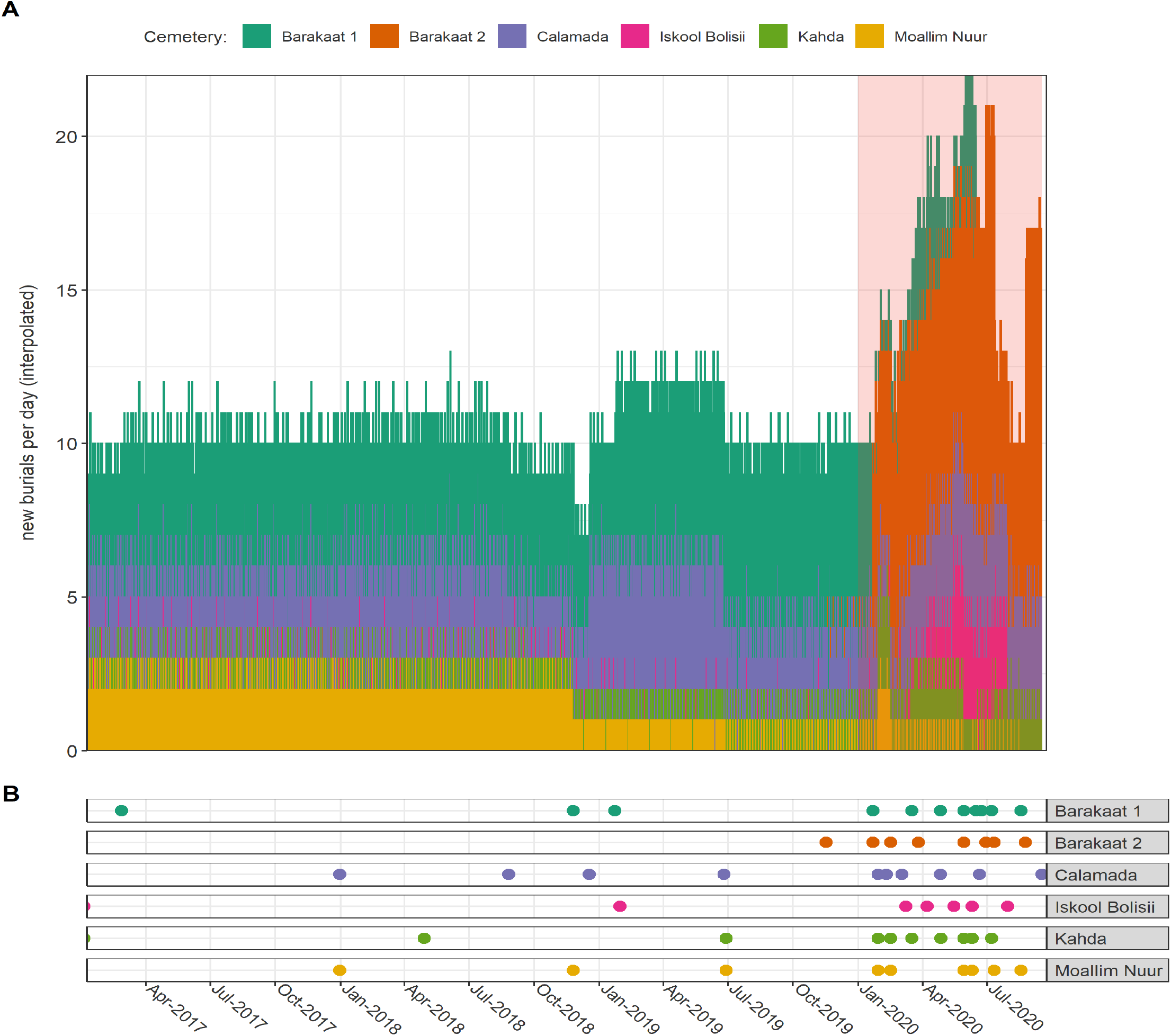
Panel A: Interpolated time series of daily burials, as estimated from satellite imagery, by cemetery. Panel B: Dates for which satellite imagery was acquired and analysed, by cemetery.

When combined with alternative population denominators, the above estimates yielded daily burial rates per 10,000 population of around 0.06 to 0.08 in January 2017, depending on the population source, declining to some 0.04 to 0.05 by the end of 2019 (Figure 6). Burial rates increased during 2020 (Figure 7), with a ratio to pre-pandemic levels averaging 1.5-fold and peaking at 2.2-fold.

**Figure 6.**
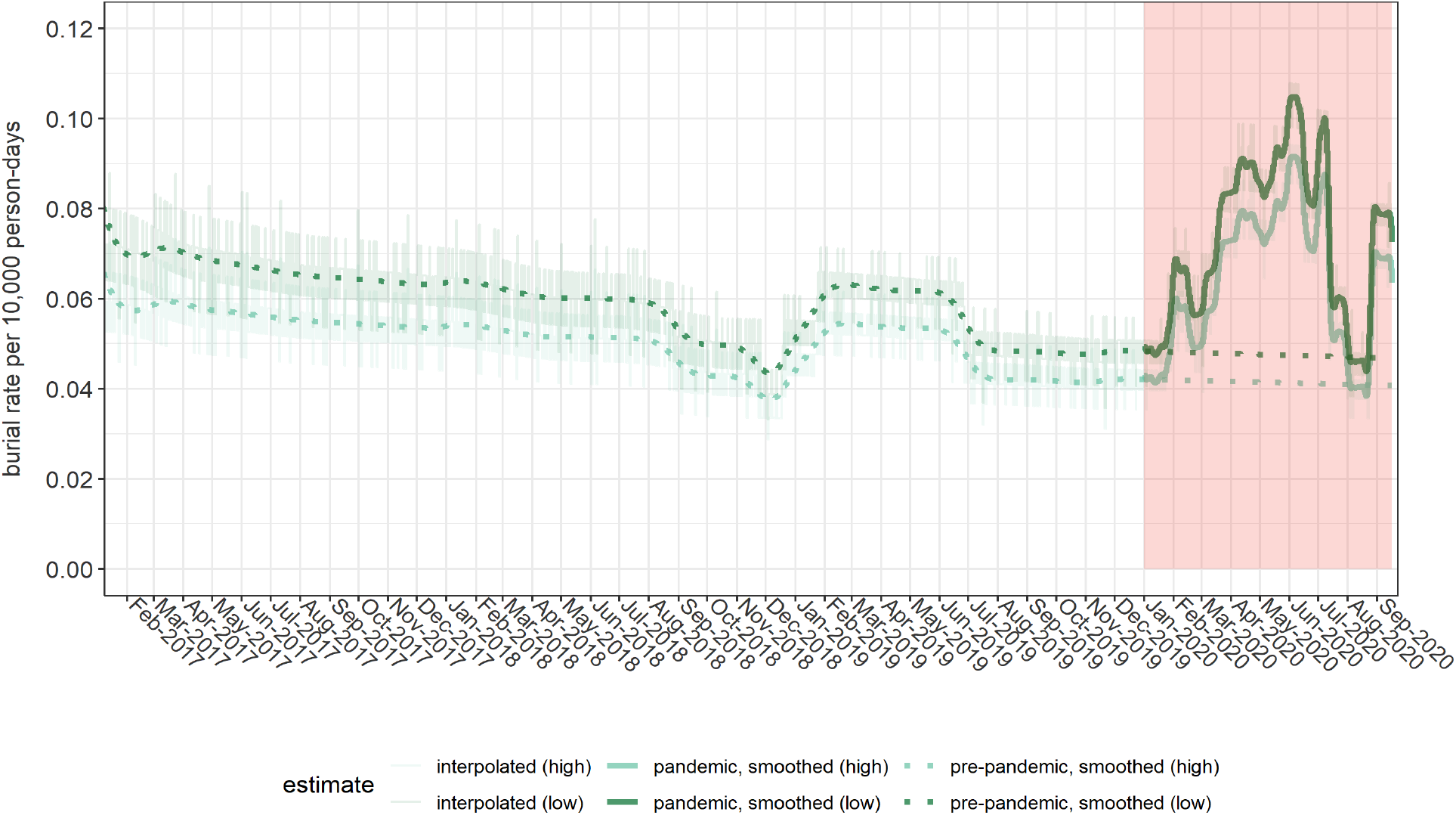
Interpolated and smoothed burial rates per 10,000 person-days across the six cemeteries analysed. Here, ‘high’ and ‘low’ denote alternative estimates computed using the higher (WorldPop 2015) and lower (WorldPop 2020) of the two population denominator scenarios. The pre-pandemic smoothed trend is projected into 2020 to provide a counterfactual level in the absence of COVID-19 and related disruptions.

**Figure 7.**
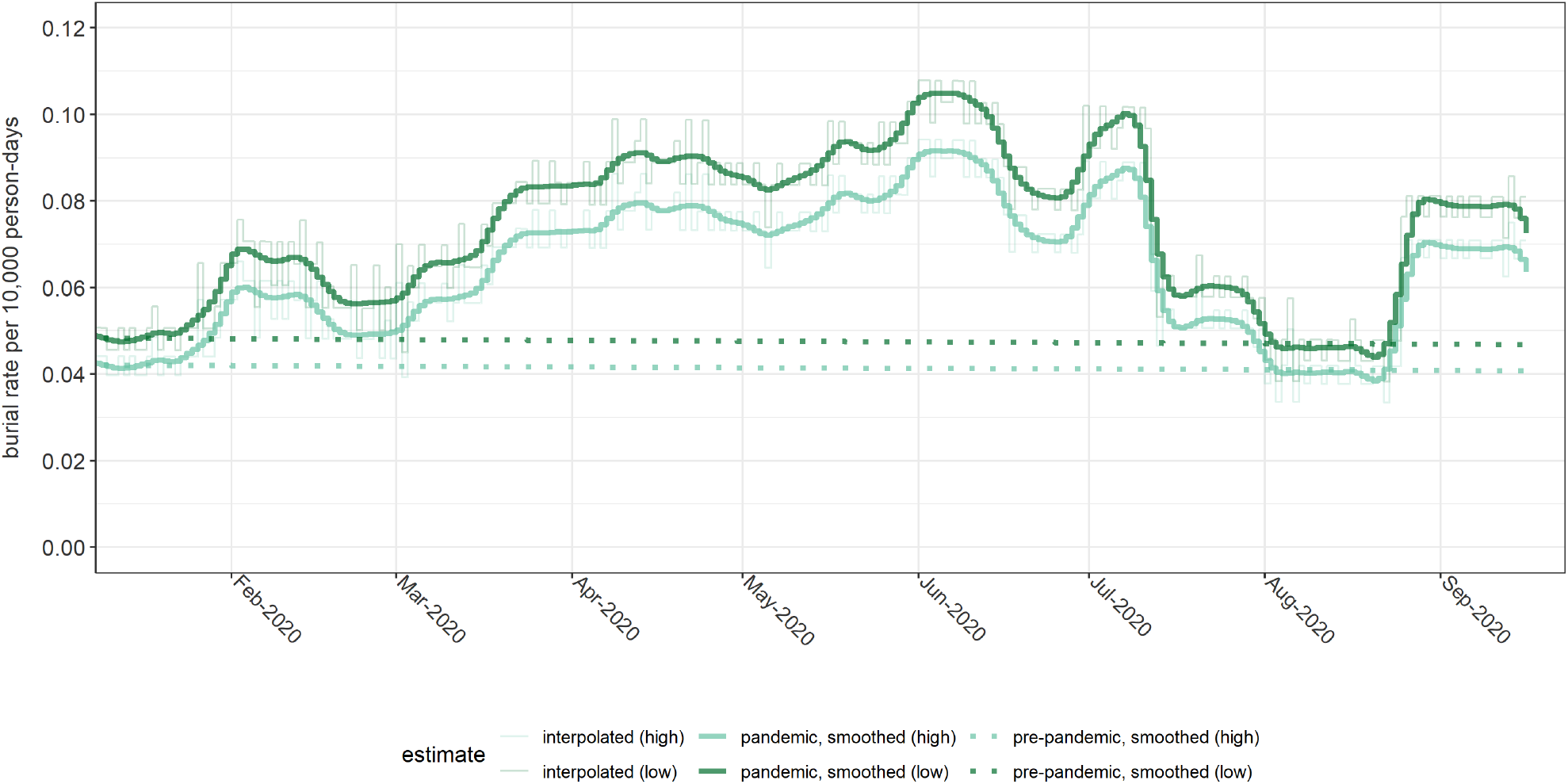
Interpolated and smoothed burial rates per 10,000 person-days across the six cemeteries analysed, during 2020.

Even before the pandemic, our estimated burial rates were about one third to one-tenth of the range in crude death rate (CDR) estimated by previous surveys in Banadir (Table 1). When scaled to this plausible range of baseline CDR, excess death toll between January and September 2020 ranges between 3,200 and 11,800 depending on the assumptions used (Table 3).

**Table 3.** Estimates of the total, counterfactual (no-pandemic) and excess death tolls in Banadir Region, Jan-Sep 2020, by population estimate, used and assumed baseline crude death rate.

| Baseline CDR†<br>(as of 1 Jan 2017) | Estimated death toll |  |  |
| --- | --- | --- | --- |
|  | Total | Baseline<br>(counterfactual) | Excess |
| 'High' population scenario (WorldPop 2015) |  |  |  |
| 0.20 | 11,200 | 7,300 | 3,900 |
| 0.25 | 14,000 | 9,100 | 4,900 |
| 0.30 | 16,800 | 10,900 | 5,900 |
| 0.35 | 19,600 | 12,700 | 6,900 |
| 0.40 | 22,400 | 14,500 | 7,900 |
| 0.45 | 25,200 | 16,300 | 8,900 |
| 0.50 | 28,000 | 18,100 | 9,900 |
| 0.55 | 30,800 | 20,000 | 10,900 |
| 0.60 | 33,600 | 21,800 | 11,800 |
| 'Low' population scenario (WorldPop 2020) |  |  |  |
| 0.20 | 9,200 | 6,000 | 3,200 |
| 0.25 | 11,500 | 7,500 | 4,000 |
| 0.30 | 13,800 | 9,000 | 4,900 |
| 0.35 | 16,100 | 10,500 | 5,700 |
| 0.40 | 18,400 | 12,000 | 6,500 |
| 0.45 | 20,800 | 13,500 | 7,300 |
| 0.50 | 23,100 | 15,000 | 8,100 |
| 0.55 | 25,400 | 16,500 | 8,900 |
| 0.60 | 27,700 | 18,000 | 9,700 |

† Crude death rate per 10,000 person-days.

**Table 4.** Comparison of alternative burial sources, for months with sufficient data.

| Cemetery | Year | Month | Satellite imagery | Barakaat Development Committee |
| --- | --- | --- | --- | --- |
| Barakaat 1 + 2 | 2019 | 4 | 155 | 494 |
| Barakaat 1 + 2 | 2019 | 5 | 160 | 532 |
| Barakaat 1 + 2 | 2019 | 6 | 155 | 473 |
| Barakaat 1 + 2 | 2020 | 1 | 178 | 778 |
| Barakaat 1 + 2 | 2020 | 2 | 209 | 389 |
| Barakaat 1 + 2 | 2020 | 3 | 279 | 390 |
| Barakaat 1 + 2 | 2020 | 4 | 313 | 609 |

### Comparison with other available sources

When compared with figures provided by Barakaat Cemetery Development Committee, our estimates were substantially lower.

### Key informant interviews

Informants stated that government imposed COVID-19 health-related measures were not effectively observed. In March 2020, authorities declared a lockdown and imposed restrictions on public movements and meetings including closure of schools, government offices and restriction of international travel. In reality community interactions and public gatherings continued to function as normal as hotel, teashops, mosques, and other public places remained open.

Informants reported an increase in the number of deaths in the month of Ramadan (April-May). A few respondents reported some of these deaths as being caused by a flu-like disease. Informants also identified a capacity gap among health care professionals, the inadequacy of health equipment such as ventilators, personal protective equipment, and other rudimentary equipment. “Shortage of ICU beds contributed to deaths as a majority of the dead died of breathing difficulties. Imagine less than 20 ICU beds are available in Mogadishu where almost 4 million persons live” (local health professional, November 2020). Informants also mentioned elevated prices of face masks and antiseptics as an access barrier the population, potentially hampering the containment of COVID-19.

The responses from the key informants appear to point towards an overburdened public health system struggling to deal with the increasing severity of the epidemic. This is consistent with the evolution in burial activity over the analysis period shown in Figure 5, especially Barakaat 1 and 2 cemeteries. “In May this year, we have seen a substantial increase in the number of deaths almost we have seen 600 people in one month. We’ve never seen such increase in the number of deaths” (BCDC member, November 2020). Informants also highlighted the stigma surrounding COVID-19 and its role in driving people to seek care at home rather than hospitals. “Families refused to take their ailing patients to the de Martini hospital fearing that they will not be able to see them again once admitted” (local health professional, November 2020). Lastly, respondents reported lack of appropriate COVID-19 burial protocols at funerals.

## Discussion

Given the well-characterized delay from infection to death(Baud et al., 2020), the peak of COVID-19 deaths reported by this study suggests a much earlier introduction into Somalia than hitherto believed, possibly as far back as December 2019. During this peak, anecdotal reports from media and government authorities suggest a substantial increase in burial rates(BBC News, 2020; Jason Burke and Abdalle Ahmed Mummin, 2020). Our estimates are broadly in line with initial modelling predications(Projections of COVID-19 epidemics in LMIC countries | CMMID Repository, n.d.). While much of the excess death toll is likely to be COVID-19 cases, some may be attributable to the indirect effects of the pandemic, e.g., socio-economic disruptions or reduced access to health services due to social distancing restrictions and overwhelmed or repurposed health facilities(Roberton et al., 2020). This has been suggested by research in the COVID-19 pandemic(Giovanni Forchini, Alessandra Lochen, Timothy Hallett, Paul Aylin, Peter J. White, Christl Donnelly, Azra Ghani, Neil Ferguson, 2020) and has also been documented during the West Africa Ebola epidemic (Elston et al., 2017). Scepticism about the existence of COVID-19 as well as the lack of social distancing(Shahow, 2021) and the potential for funerals as superspreading may have contributed to the higher transmission of the virus and could partly explain the scale of excess deaths. Knowledge on preventative measures that may have mitigated the impact of the pandemic has been found to be low among the general population as well as IDPs in Mogadishu.(Alawa et al., 2020) Additionally, fear and stigma associated with hospital care and an existing preference for homecare(Tanne et al., 2020) could both have increased the proportion of unreported deaths and burials. As with a similar study in Yemen, results indicate a considerable under ascertainment(Besson et al., 2021) of excess deaths. Given the scarcity of COVID-19 testing in Somalia, such an estimate provides a useful metric for establishing the full impact of the pandemic. Ultimately, a long-term COVID-19 control strategy for Somalia needs to be predicated upon sound public health principles: renewed lockdowns and harmful socio-economic restrictions may not be acceptable, and their health impact is increasingly documented. Community-led, culturally appropriate forms of mitigation may, however, warrant consideration (UNICEF et al., 2021): in particular, local approaches to shield the elderly and vulnerable for a limited period during COVID-19 waves could be adopted by people, with support from humanitarian and development actors(Dahab et al., 2020; van Zandvoort et al., 2020). Social awareness and preventive measures should be prioritized as the weak health system in Somalia, cannot provide services to the severe cases of COVID-19. Such strategies should be combined with a pragmatic approach to surveillance, utilising not just formal testing but also symptom monitoring (e.g. through social media or sentinel surveillance in health facilities) as well as mortality data collection to rapidly detect and assess the severity of renewed COVID-19 waves, whenever these do occur.

### Limitations

Systematic or random error may have arisen from the method for imputing missing grave counts. Extrapolation from limited area samples to fill in some of the starting values likely increased error. Moreover, the model to impute missing graves had moderate precision, likely due to the low number of observations used to train it. Further error may have arisen from problems with counting individual graves in satellite images. The above sources of error arise from different statistical processes and are difficult to combine into a single estimation framework: as such, we were unable to produce realistic confidence intervals for the estimates. Overall, there was strong evidence that our estimates of burials per population were a considerable under-estimate of the plausible death rate in the Banadir region: this may have been due to insensitivity of imagery analysis, failure to comprehensively identify and analyse all burial sites used by the population, decedents being buried in their communities of origin outside Banadir region, and/or burials taking place in informal plots (e.g. nearby private residences). Scaling the baseline burial rate to a more plausible CDR level rests on a strong assumption that the under-estimation bias of our method remains approximately constant over time. Satellite imagery analysis was undertaken retrospectively, meaning there were constraints with the data available being limited to what had already been sporadically collected by VHR satellite imagery providers. Additionally, budgetary limitations meant acquiring new data on demand at 30cm for Banadir was not possible, leading to reliance on archive data, limiting the number of valid data points we were able to collect. Undertaking similar studies on a proactive basis would facilitate the collection of VHR in a timely manner at a cadence which would allow for greater density of data points both in space and time. Lastly, the detection algorithm developed cannot detect graves in the areas where the ground has degraded due to erosion etc. or areas where tree growth has obscured the grave plots.

## Conclusions

The findings from this study suggest substantial underreporting of COVID-19 in Mogadishu, Somalia. As with our previous work in Aden governorate, Yemen, we believe the satellite imagery method coupled with local verification and qualitative data is a promising epidemiological tool for assessing excess mortality in resource-constrained and fragile/crisis-affected settings. Our study suggests that the Somali population is indeed highly vulnerable to COVID-19, with no evidence of reduced susceptibility after accounting for obvious age differences between Somalia and high-income countries. The current observation of a severe renewed wave suggests that existing immunity was insufficient to grant herd protection, either due to waning of immunity or the introduction of more transmissible variants. These observations underscore the importance of reinforcing control measures for COVID-19, including behaviour change and hygiene, and rolling out COVID-19 vaccination to at least highly vulnerable groups within Somalia.

## Data Availability

All data related to the statistical analysis can e accessed in the Github repository below.

https://github.com/francescochecchi/mogadishu_burial_analysi

## Ethical approval

Ethical approval was received from the ethics review committee of the London School of Hygiene & Tropical Medicine (REF: 22458) as well as the ethics review committee of the Somali Disaster Resilience Institute (REF: RB-0123)

## Acknowledgments

We would like to acknowledge Sheikh Abdullahi Ali of the Barakaat Cemetery Development Committee and Dr Mohamed Fuje of the national COVID-19 task force for their support.

## References

Alawa J, Al-Ali S, Walz LA, Wiles E, Harle N, Awale MA, et al. Knowledge and perception of COVID-19, prevalence of pre-existing conditions, and access to essential resources and health services in Somali IDP camps. MedRxiv 2020:2020.08.17.20176271. https://doi.org/10.1101/2020.08.17.20176271.

Baud D, Qi X, Nielsen-Saines K, Musso D, Pomar L, Favre G. Real estimates of mortality following COVID-19 infection. Lancet Infect Dis 2020;20:773. https://doi.org/10.1016/S1473-3099(20)30195-X.

BBC News. The gravedigger’s truth: Hidden coronavirus deaths. Br Broadcast Corp News Africa 2020.

Besson EK, Norris A, Bin Ghouth AS, Freemantle T, Alhaffar M, Vazquez Y, et al. Excess mortality during the COVID-19 pandemic in Aden governorate, Yemen: A geospatial and statistical analysis. MedRxiv 2020. https://doi.org/10.1101/2020.10.27.20216366.

Besson ESK, Norris A, Ghouth ASB, Freemantle T, Alhaffar M, Vazquez Y, et al. Excess mortality during the COVID-19 pandemic: A geospatial and statistical analysis in Aden governorate, Yemen. BMJ Glob Heal 2021;6:4564. https://doi.org/10.1136/bmjgh-2020-004564.

Bryman A. Social Research Methods. Oxford: 2012.

Centre national d’études spatiales. Orfeo ToolBox – Orfeo ToolBox is not a black box 2019. https://www.orfeo-toolbox.org/ (accessed May 6, 2021).

Checchi F. GitHub - francescochecchi/mogadishu_burial_analysis: R scripts and dataset for analysis of burial patterns in Mogadishu, Somalia (2017-2020), as part of an effort to quantify mortality attributable to the COVID-19 pandemic. 2021. https://github.com/francescochecchi/mogadishu_burial_analysis (accessed May 10, 2021).

Cloudfactory. Image Annotation for Computer Vision: A Guide to Labeling Visual Data for Your Machine Learning Project 2020.

Dahab M, van Zandvoort K, Flasche S, Warsame A, Ratnayake R, Favas C, et al. COVID-19 control in low-income settings and displaced populations: what can realistically be done? Confl Health 2020;14:1–6. https://doi.org/10.1186/s13031-020-00296-8.

Directorate of National Statistics Federal Government of Somalia. The Somali Health and Demographic Survey 2020. 2020.

Elston JWT, Cartwright C, Ndumbi P, Wright J. The health impact of the 2014–15 Ebola outbreak. Public Health 2017;143:60–70. https://doi.org/10.1016/j.puhe.2016.10.020.

Giovanni Forchini, Alessandra Lochen, Timothy Hallett, Paul Aylin, Peter J. White, Christl Donnelly, Azra Ghani, Neil Ferguson KH. Report 28 - Excess non-COVID-19 deaths in England and Wales between 29th February and 5th June 2020. Imp Coll London 2020.

Google Earth. 2021. https://www.google.co.uk/intl/en_uk/earth/ (accessed May 6, 2021).

Google Maps. 2021. https://www.google.co.uk/maps/about/#!/ (accessed May 6, 2021).

Jason Burke, Abdalle Ahmed Mummin. Somali medics report rapid rise in deaths as Covid-19 fears grow. Guard 2020.

Linard C, Gilbert M, W. Snow R, Abdisalan M. Noor AJT. Population Distribution, Settlement Patterns and Accessibility across Africa in 2010. PLoS One 2012.

Maxar. Maxar-Securewatch 2021.

Ministry of Health Somalia. COVID-19 Situational Update. COVID-19 Situational Updat 2021. https://moh.nomadilab.org/ (accessed May 6, 2021).

OpenStreetMap. 2021. https://www.openstreetmap.org/#map=6/54.910/-3.432 (accessed May 6, 2021). Projections of COVID-19 epidemics in LMIC countries | CMMID Repository. n.d.

Roberton T, Carter ED, Chou VB, Stegmuller AR, Jackson BD, Tam Y, et al. Early estimates of the indirect effects of the COVID-19 pandemic on maternal and child mortality in low-income and middle-income countries: a modelling study. Lancet Glob Heal 2020;8:e901–8. https://doi.org/10.1016/S2214-109X(20)30229-1.

Shahow AA. The New Humanitarian | Who’s afraid of COVID-19? Somalia’s battle with the virus. New Humanit 2021. https://www.thenewhumanitarian.org/analysis/2021/5/5/whos-afraid-of-covid-19-somalias-battle-with-virus (accessed May 11, 2021).

Tanne JH, Hayasaki E, Zastrow M, Pulla P, Smith P, Rada AG. Covid-19: How doctors and healthcare systems are tackling coronavirus worldwide. BMJ 2020;368. https://doi.org/10.1136/bmj.m1090.

UNDP. Floods, locusts and COVID-19; Somalia’s triple threat - Somalia. United Nations Dev Program 2020.

UNFPA Somalia. Saving the lives of Somali mothers and newborns amid COVID-19. United Nations Popul Fund 2020.

UNHCR. SOMALIA: INTERNAL DISPLACEMENTS. 2021.n.d. https://unhcr.github.io/dataviz-somalia-prmn/index.html#reason=&month=&need=&pregion=&pdistrictmap=&cregion=&cdistrictmap=&year=2021 (accessed March 30, 2021).

UNICEF, UNFPA, WHO, SickKids’ Center for Global Child Health. Direct and indirect effects of the COVID-19 pandemic and response in South Asia. Kathmandu, Nepal: 2021.

UNSC. COVID-19, Severe Locust Outbreaks Compound Economic, Security Woes in Somalia Ahead of Long-Awaited Elections, Experts Tells Security Council | Meetings Coverage and Press Releases. United Nations Secur Counc 2020.

Wariyaha Muqdisho. Filish: Dad badan oo aan garanayo ayaa Covid-19 ugu dhintay Muqdisho. Garowe Online 2020. https://www.garoweonline.com/en/news/somalia/filish-dad-badan-oo-aan-garanayo-ayaa-covid-19-ugu-dhintay-muqdisho (accessed October 3, 2020).

WHO. Responding to COVID-19 in Somalia : Progress Report 6 months of resilience and strength. 2020. Wordpop Gridded Population Estimate Datasets and Tools. 2020.

van Zandvoort K, Jarvis CI, Pearson CAB, Davies NG, Nightingale ES, Munday JD, et al. Response strategies for COVID-19 epidemics in African settings: a mathematical modelling study. BMC Med 2020;18:324. https://doi.org/10.1186/s12916-020-01789-2.

