## Supplementary files for "Excess mortality during the COVID-19 pandemic: a geospatial and statistical analysis in Mogadishu, Somalia"

### Supplementary tables and figures

**Table S1. Predictive GAMLSS model to impute new graves.**

| Term | Rate ratio | p-value |
|---|---|---|
| Penalised B-spline smoothing terms for the mean (expectation) parameter): | | |
| Surface area growth ($m^2$) | 0.882 | < 0.001 |
| Penalised B-spline smoothing terms for the shape (over dispersion) parameter): | | |
| Surface area growth ($m^2$) | -0.0003 | 0.910 |
| | Akaike Information Criterion = 336.6 | Residual degrees of freedom = 17.2 |

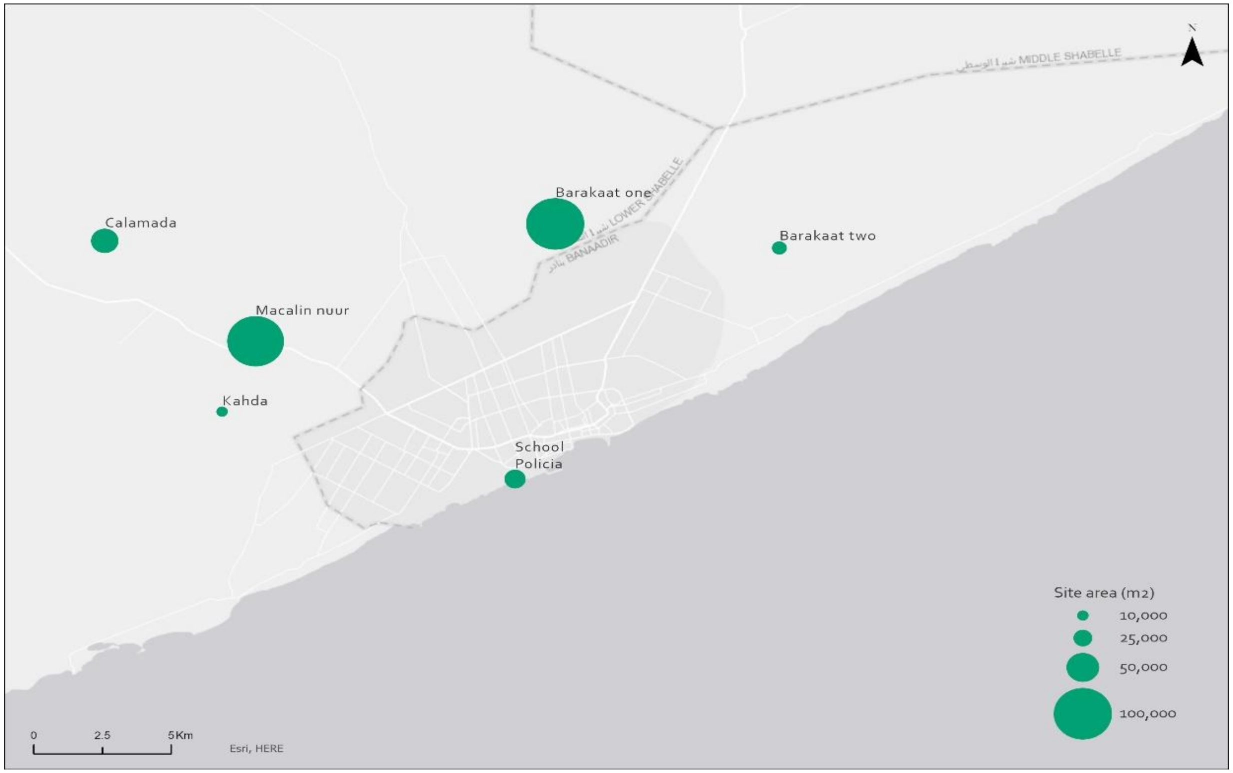

**Figure S1. Analysed cemeteries by size**

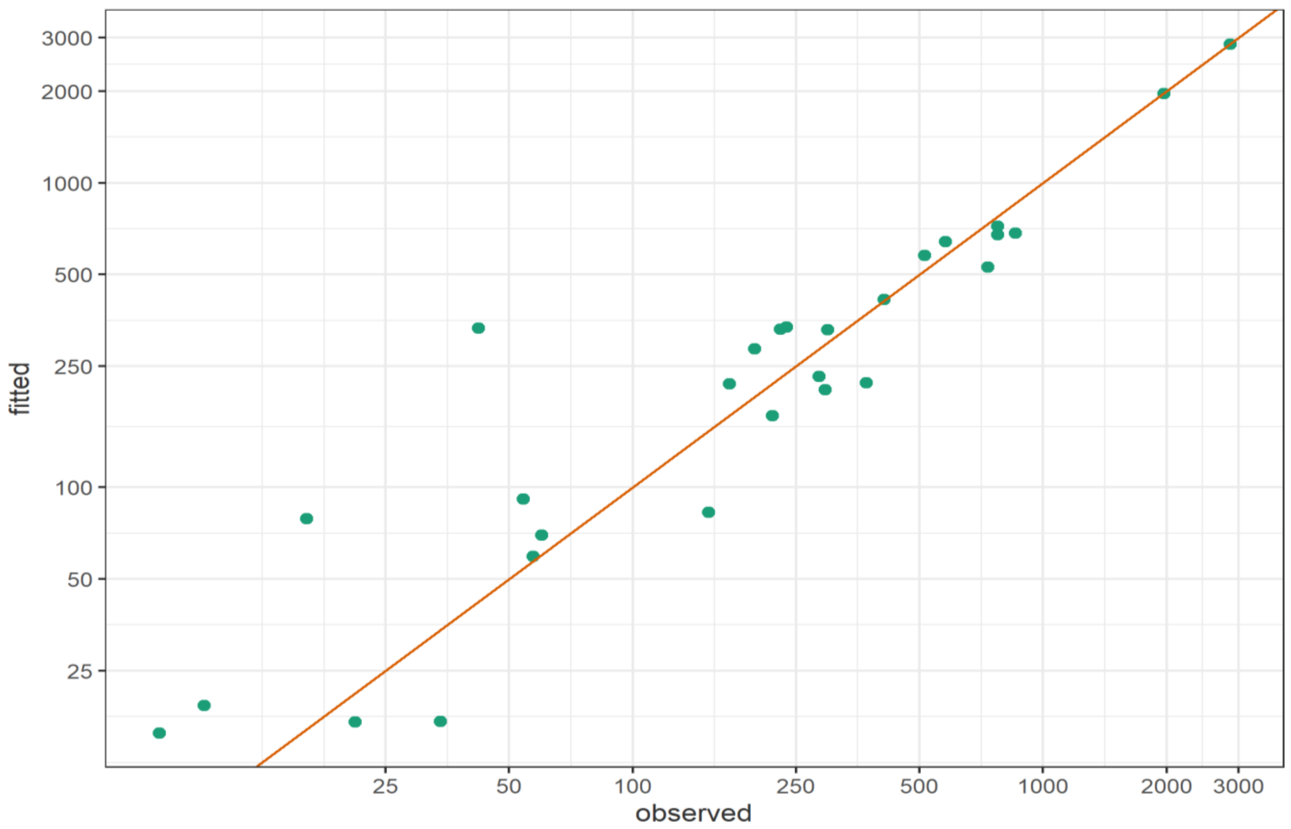

**Figure S2. Fitted versus observed number of new graves. The red line indicates perfect agreement between observations and predictions.**

#### Field photos

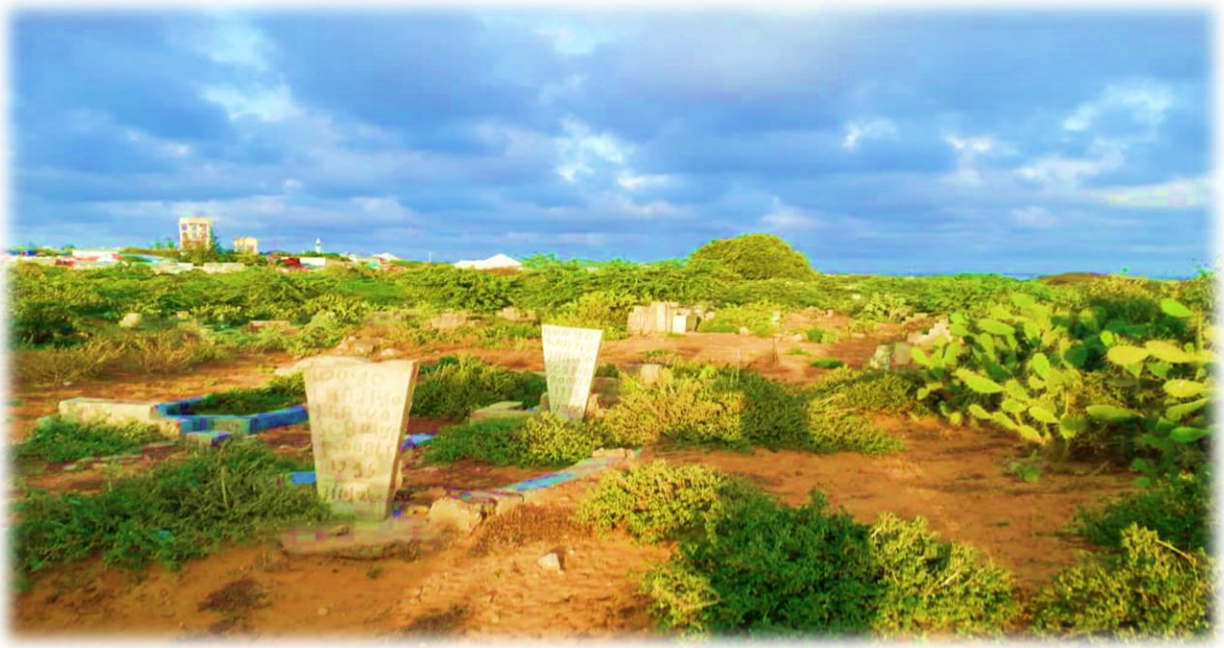

**Figure S3. Moalim Nuur Cemetery**

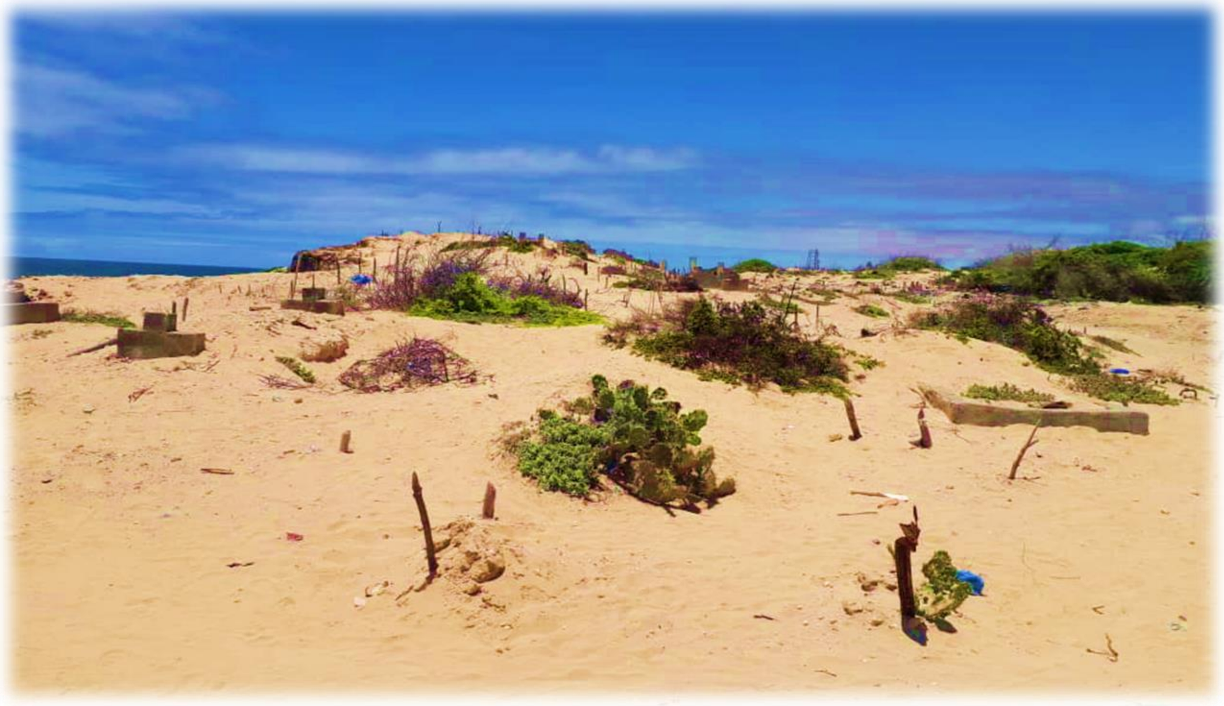

**Figure S4. School Policio Cemetery**

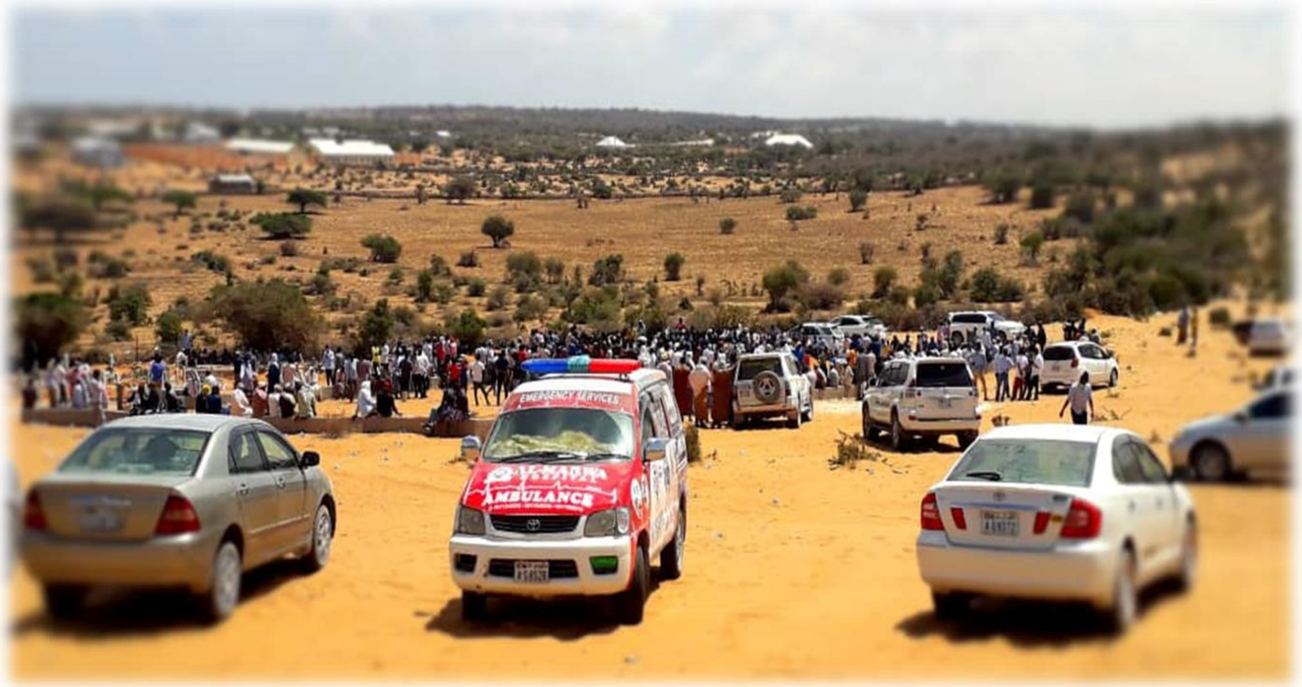

**Figure 1 Burial gatherings during the COVID19. When person dies regardless of the cause people gather and pray for the deceased.**

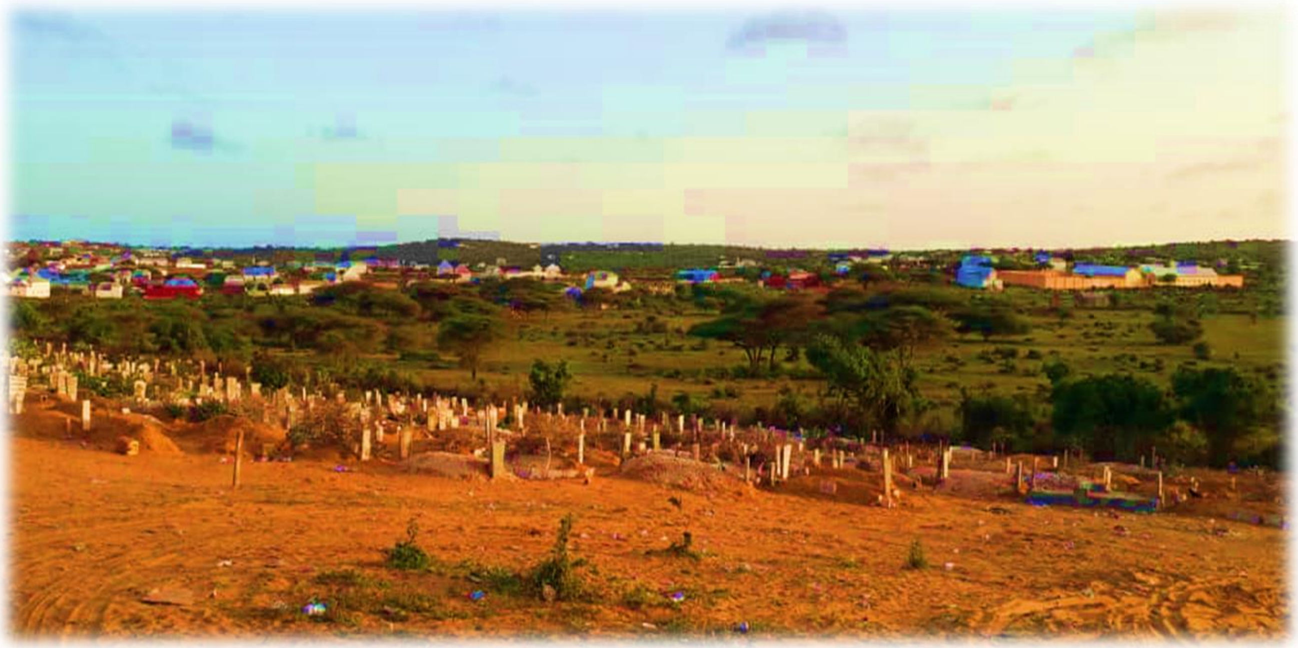

**Figure S6. Kaxda Cemetery**

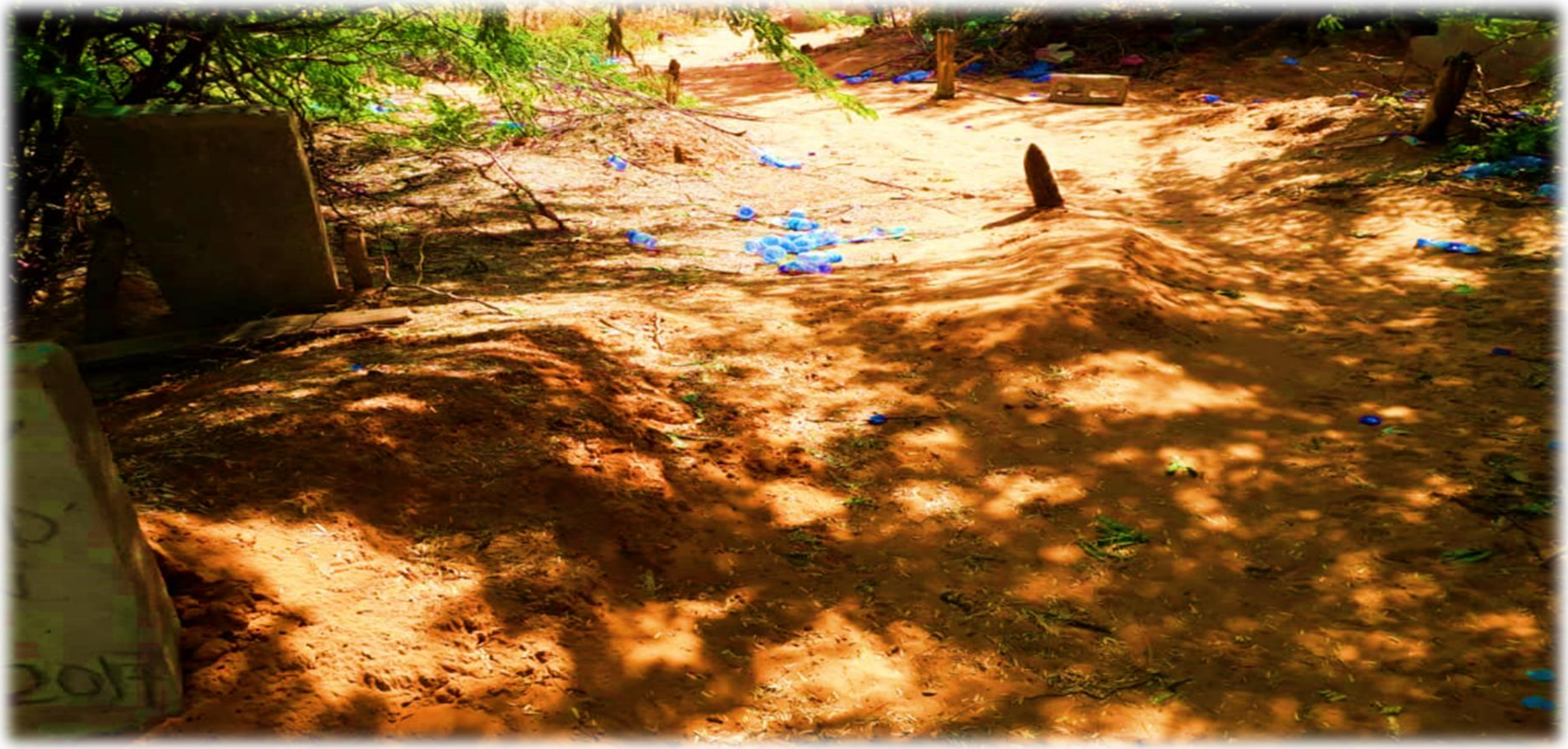

**Figure S7. A group of graves covered by trees in Abay Dhahan Cemetery**

**Table S2. Cemetery Profile**

| No | Name | District | Ownership | Status | Details |
|---|---|---|---|---|---|
| I. | Barakaat one (Haji Abdi) | Yaqshid | Private | Closed | The old Barakaat Cemetery was at the Mogadishu University's main campus, outside of Mogadishu. Established 20 years ago and it consisted of 550 plots of land. It is full and there are no official burials except some special conditions like senior government members, religious scholars, or infants. Wahar-Ade (Barakaat two) is the main burial that replaced the first Barakaat site. This site has been abandoned one year ago but is used as a reference. It was the main burial site for Mogadishu. |
| II. | Barakaat two | Heliwaa | Private | Functional | Wahar Cade(Barakaat Two) cemetery is in Waharadde, Heliwaa District, and consists of 370 plots of lands. It has been functioning for around a year starting from March 2020. It serves all the city districts as documented. It is also the sole site with numbering and order. Other locations don't have proper numbering and management, but this field is managed by private individuals. |
| III. | Calamada, The third Barakaat Cemetery | Afgoye-Lowershabele-Mogadishu's Outskirts | | | The site locates in between Eelash and Afgooye (Calamada); this cemetery consists of 300 plots of land. It was established in 2009 and is currently functional although it is not used as much as the other sites. People from Afgooye (a city in Lower Shabelle, 30KM away from Mogadishu), Medina and its surrounding areas use this site to bury their dead loved ones. |
| IV. | Kahda Cemetery | Kaxda | Public | Functional | Kaxda has significant land for burials but it didn't provide numbers for reference. Some of the locals told us that the site holds around 2000 to 3000 graves. It has been functional for more than ten years. During the COVID-19 pandemic, the frequency of burials increased. It was more than two folds for instance during the emergency. The site recorded more than 30 burials in a single month which is more than double of the norm. |
| V. | Abaay Dhaxan Cemetery | Wadajir | Private | Functional | It doesn't have numbering and order. It is usually used by the poor communities. It also the main burial for specific clans (minorities). |
| VI. | Jazeera | Wadajir | Private | Functional | This site is functioning. It has quite a few algarroba tree. |
| VII. | Elade (Old Grave) | Kaaraan | Private | Closed | Elade located outskirts of Mogadishu and is used by few people. The area around El Ade has different small cemeteries. Most of the locals use the newly established Barakaat facility in northern Mogadishu. |

| | | | | | |
|---|---|---|---|---|---|
| VIII. | Macalin nuur (Moallim Nurr) | Deynile( Garasbaley) | Private | Officially Closed | It was one of the main graveyards in Mogadishu, but it has not been functional in recent years. Earlier, IDP resettled on the graves. Parts of the graveyard were wiped out. But, still, some informal burials are undertaken. Calamada site services the locality it used to serve. |
| IX. | School Policia | X/Jajab | Veteran/National | Functional | School Policio site is mainly used by the poor and IDPs. It is not organized rather it has been overused and overcrowded. It is located near the ocean. It doesn't have a management team. |
| X. | Madiina Hospital Cemetery | Wadajir | National | Not-Functional | Usually, the bodies of the wounded individuals were buried; it was a piece of land owned by the Madina Police Hospital. |
| XI. | Abdirashid Cemetery | Hodan | Public | Not-Functional | Abdirashid has been abandoned more than a decade ago and there is no indication of recent activity. Some of the cemetery land was occupied for residential proposes. |
| XII. | Jeneral Dauud | Wardhigley | Military | Not-functional | One of the old cemeteries in Mogadishu. It is no longer functional and useful. |
| XIII. | Adan Adde | Yaqshiid | Private-Family | Private-Family owned | Burial of elites including the first president and second prime minister of Somalia. |
| XIV. | Deynile-Sites(Banglo, Zeynab Qoobey cemeteries) | | | Private-Family | But, most of the people go to the Barakaat cemetery to avoid digging and self-service as the BCDO facilitates the burial services. Family-Sub clan cemeteries don't have management, and your relatives must dig the burial hall instead of being served. |
